# Convergent validity of a person-dependent definition of a low back pain flare

**DOI:** 10.1101/2025.01.11.25320390

**Authors:** Pradeep Suri, Anna Korpak, Andrew Timmons, Adrienne D. Tanus, Hannah Brubeck, Nathalia Costa, Carina A. Staab, Paul Hodges, Clinton J. Daniels, Patrick J. Heagerty, Mark P. Jensen

## Abstract

Exacerbations of existing low back pain (LBP) or new LBP episodes are colloquially referred to as “flares”. Although the experience of flares is common to many people with LBP, few validated measures enable people to self-report if they are experiencing a flare. This study examined the convergent validity of a person-dependent definition of flares (“a worsening of your low back pain that lasts from hours to weeks”) as compared to (1) LBP intensity, (2) LBP-related pain interference, and (3) analgesic use, in a large, prospective research study of Veterans with LBP. Veterans seeking care for LBP (n=465) were followed prospectively for up to 1 year. Participants completed up to 36 scheduled surveys and additional patient-initiated surveys (triggered by the onset of new flares) over follow-up. Each survey inquired about current flare status, pain intensity measured on a 0 to 10 numeric rating scale (NRS), LBP-related pain interference, and analgesic use. Linear mixed-effects models estimated the association between current flare status and pain intensity, with and without adjustment for potential confounding factors; secondary analyses examined associations with pain interference and analgesic use. In longitudinal analyses of 11,817 surveys, flare status was significantly associated with a 2.8-NRS-point greater pain intensity (p<0.0001), with and without adjustment for other factors. Statistically significant associations were found between flare status and LBP-related pain interference and analgesic use. New flare periods were associated with impacts on coping, functional limitations, and mood/emotions. These findings support the convergent validity of a person-dependent flare definition.

**SUMMARY:** A person-dependent definition of “flare” among individuals with low back pain has convergent validity as shown by its associations with pain intensity and pain interference.

## 1. INTRODUCTION

Low back pain (LBP) is the leading cause of years lived with disability worldwide[7] and associated with greater healthcare costs in the United States (US) than any other health condition.[6] While low back pain has traditionally been characterized as a dichotomy of “acute” (short-duration pain) and “chronic” (long-duration pain, typically ≥3 months), it is now recognized that these features are insufficient by themselves to describe the varied trajectories and patterns of changes in pain that are part of the LBP experience.[9] Exacerbations of existing LBP or new LBP episodes, collectively referred to as “flares” or “flare-ups”, are an important feature of the LBP experience not captured by a “acute” and “chronic” dichotomy of pain. As compared to those with LBP who do not experience intermittent flares, people with LBP who do have flares tend to experience greater back pain-related disability, lost work time, and societal costs.[13] Accordingly, the prevention and treatment of LBP flares have begun to receive attention as potential ways to mitigate the substantial negative societal impact of LBP.

Table 1 presents definitions of LBP flares used in prior research on this topic. Typically, flare definitions are presented to research participants, who then endorse whether a flare is present currently and specify other aspects of flare history. Used in this manner, a flare definition is a critical part of a *measure* that can ascertain the presence or absence of a flare. Early definitions of flares focused on pain severity or intensity when presenting the flare concept to participants.[17; 18; 23] More recently, researchers have tried to establish new LBP flare definitions that incorporate participants’ perspectives on the experience of flares (Table 1) by defining flares not only in terms of increased pain but also in terms of participants’ perceptions of their reactions to pain and/or the effects of flares on pain-related outcomes or “domains” other than pain intensity, such as mood and coping.[1] An alternative to these types of researcher-specified flare definitions is to allow participants to decide what a flare is from their perspective: a *person-dependent flare definition*. This flare definition can then be followed by additional question items that allow participants to specify the pain-related *domains* involved in a given flare. To date, few flare definitions [3; 17] [4] have been subjected to quantitative assessments of convergent validity with regards to pain intensity and/or other factors associated with the pain experience (Table 1).

**Table 1.**
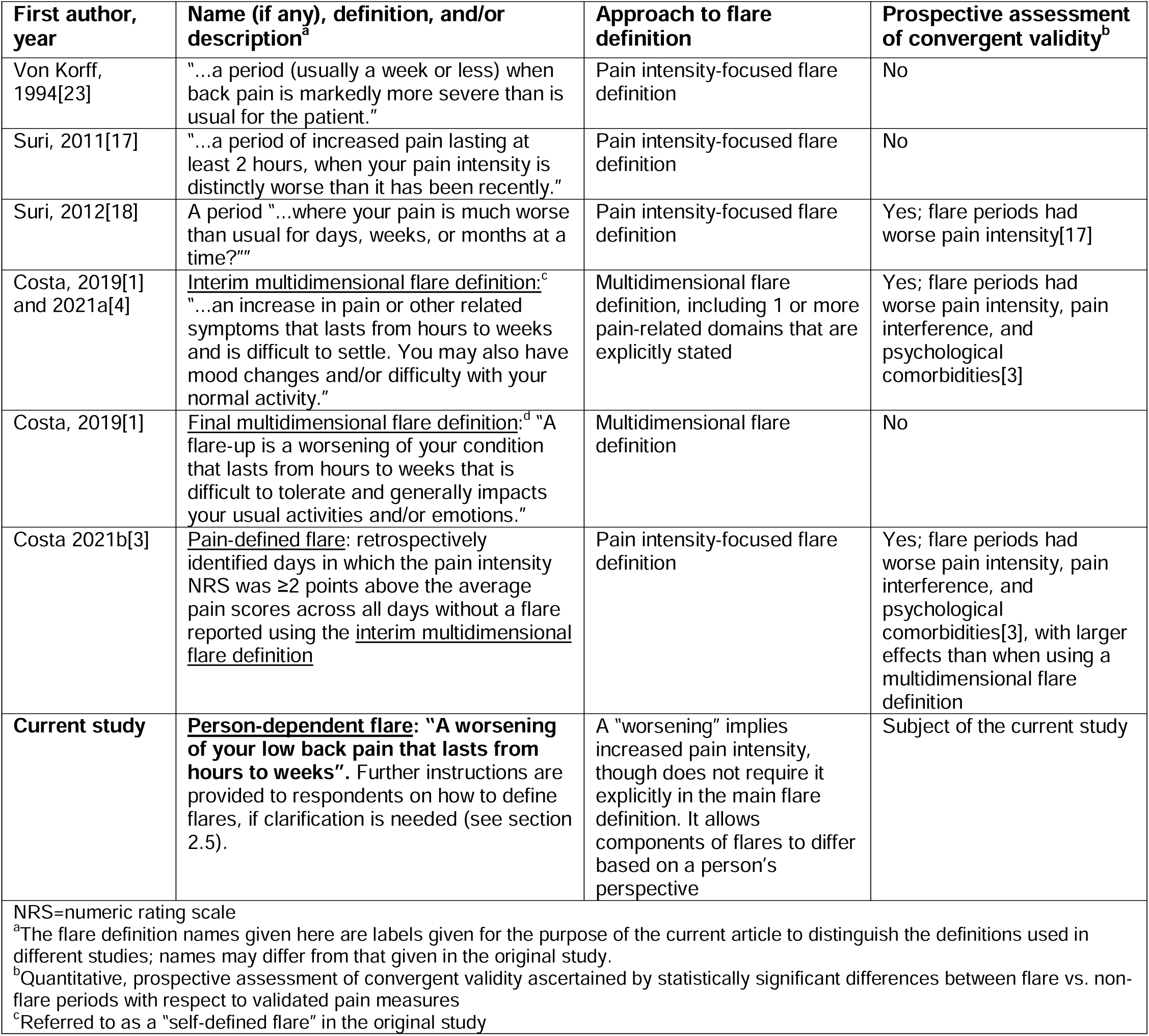
Flare definitions used in prior low back pain (LBP) research.

| First author, year | Name (if any), definition, and/or description<sup>a</sup> | Approach to flare definition | Prospective assessment of convergent validity<sup>b</sup> |
| --- | --- | --- | --- |
| Von Korff, 1994[23] | "...a period (usually a week or less) when back pain is markedly more severe than is usual for the patient." | Pain intensity-focused flare definition | No |
| Suri, 2011[17] | "...a period of increased pain lasting at least 2 hours, when your pain intensity is distinctly worse than it has been recently." | Pain intensity-focused flare definition | No |
| Suri, 2012[18] | A period "...where your pain is much worse than usual for days, weeks, or months at a time?" | Pain intensity-focused flare definition | Yes; flare periods had worse pain intensity[17] |
| Costa, 2019[1] and 2021a[4] | <u>Interim multidimensional flare definition:</u><sup>c</sup> "...an increase in pain or other related symptoms that lasts from hours to weeks and is difficult to settle. You may also have mood changes and/or difficulty with your normal activity." | Multidimensional flare definition, including 1 or more pain-related domains that are explicitly stated | Yes; flare periods had worse pain intensity, pain interference, and psychological comorbidities[3] |
| Costa, 2019[1] | <u>Final multidimensional flare definition:</u><sup>d</sup> "A flare-up is a worsening of your condition that lasts from hours to weeks that is difficult to tolerate and generally impacts your usual activities and/or emotions." | Multidimensional flare definition | No |
| Costa 2021b[3] | <u>Pain-defined flare:</u> retrospectively identified days in which the pain intensity NRS was $\geq 2$ points above the average pain scores across all days without a flare reported using the <u>interim multidimensional flare definition</u> | Pain intensity-focused flare definition | Yes; flare periods had worse pain intensity, pain interference, and psychological comorbidities[3], with larger effects than when using a multidimensional flare definition |
| **Current study** | **<u>Person-dependent flare:</u>** "A worsening of your low back pain that lasts from hours to weeks". Further instructions are provided to respondents on how to define flares, if clarification is needed (see section 2.5). | A "worsening" implies increased pain intensity, though does not require it explicitly in the main flare definition. It allows components of flares to differ based on a person's perspective | Subject of the current study |

NRS=numeric rating scale
<sup>a</sup>The flare definition names given here are labels given for the purpose of the current article to distinguish the definitions used in different studies; names may differ from that given in the original study.
<sup>b</sup>Quantitative, prospective assessment of convergent validity ascertained by statistically significant differences between flare vs. non-flare periods with respect to validated pain measures
<sup>c</sup>Referred to as a "self-defined flare" in the original study

This study examined the convergent validity of a person-dependent LBP flare definition focused on “worsening” of LBP as perceived by the participant. The study aim was to examine the validity of this flare definition as compared to outcome domains that are commonly used in LBP research, including (1) pain intensity, (2) pain interference (pain-related functional limitations), and (3) analgesic use as a reflection of the need for concurrent pain treatment. We hypothesized that, if valid, the presence of a flare using a person-dependent LBP flare definition would be significantly associated with measures of pain intensity, pain interference (pain-related functional limitations), and analgesic use.

## 2. METHODS

### 2.1 Conceptual overview

Although the concept of a “flare” is common to many pain and rheumatologic conditions,[2] there is no widespread agreement on how to operationalize the assessment of flares or what participant-reported measure should be used to ascertain whether a flare is present. In general, research has taken two approaches to LBP flare definitions.

#### 2.1.1. Pain intensity-focused flare definitions

The first approach, articulated in the earliest publications on “flares” or “flare-ups,”[18; 23] was to define flares primarily as increases in pain intensity or severity (Table 1), which we refer to henceforth as *pain intensity-focused flare definitions*. Figure 1A shows a conceptual model depicting one example of a pain intensity-focused flare definition, illustrating how a risk factor may cause a pain intensity-focused flare, which in turn might cause other downstream sequelae such as changes in mood or functional limitations (which are associated with each other). A variation on model A could include bidirectional arrows between incident LBP/increased LBP intensity and changes in mood and/or functional limitations. Numerous other models with other consequences or correlates of pain intensity-focused flares can be envisioned.

**Figure 1.**
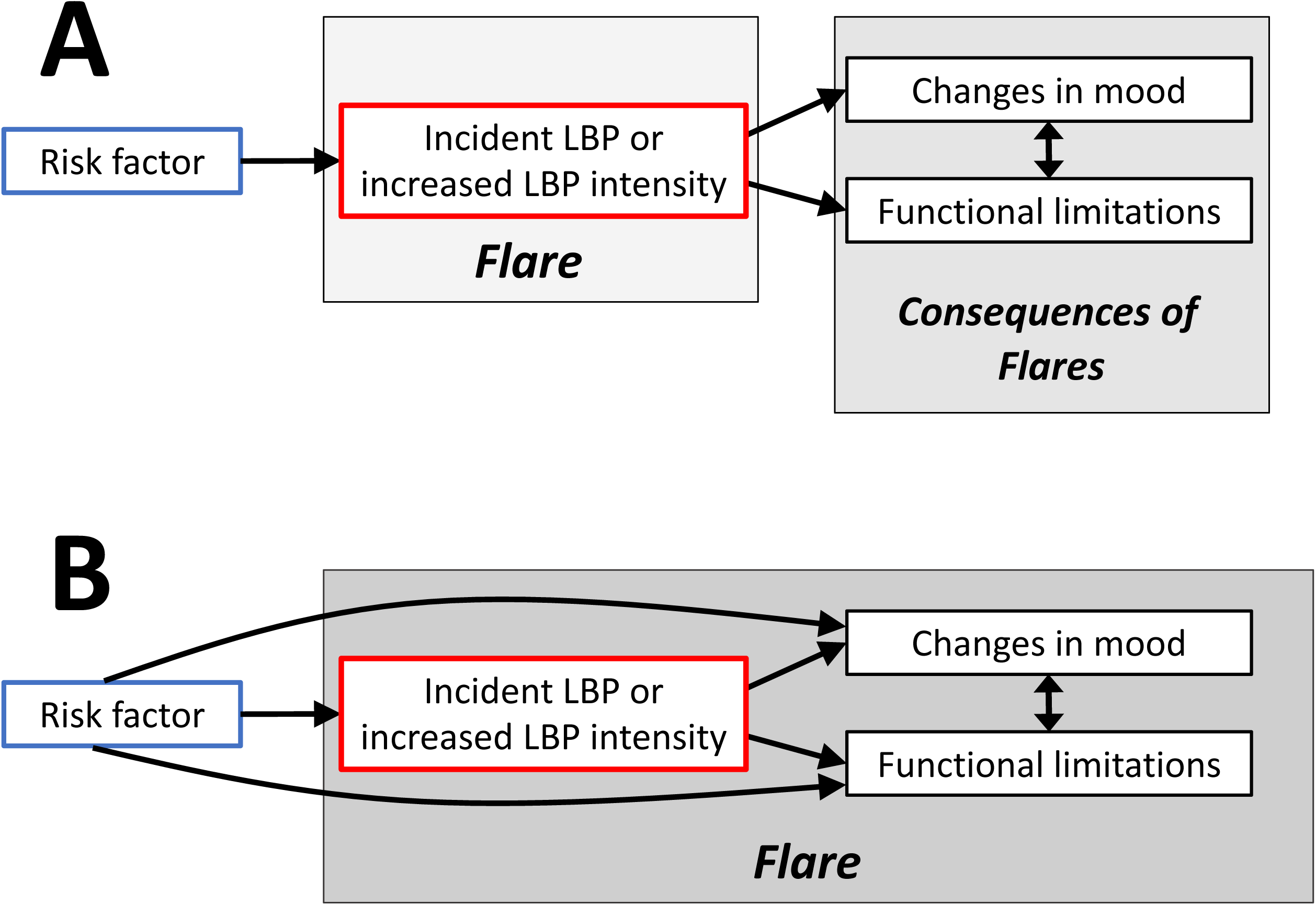
Conceptual model for how the relationship between a risk factor and flares may differ depending on the flare definition used. Figure 1A shows a conceptual model depicting a pain intensity-focused flare definition, illustrating how a risk factor may cause a pain intensity-focused flare, which in turn might cause other downstream sequelae such as changes in mood or functional limitations (which are associated with each other). Figure 1B illustrates a conceptual model exemplifying a multidimensional flare definition, in which flares are defined not only by increases in pain intensity, but also or instead are defined by changes in mood and/or functional limitations.

#### 2.1.2. Multidimensional flare definitions

The second approach has been to consider a more holistic concept of flares, defined not only by pain intensity but by other domains that are part of the pain experience, some of which may occur as a consequence of increased pain intensity and some which might occur independent of or without increases in pain intensity. We refer to this second approach as a *multidimensional flare definition*. Figure 1B illustrates a conceptual model exemplifying a multidimensional flare definition, in which flares are defined not only by increases in pain intensity, but also or instead are defined by changes in mood and/or functional limitations.

Beginning with results from an Australian online qualitative study of people with LBP by Setchell et al. [15], multidimensional flare definitions were developed by Costa and Hodges and colleagues in a multistage process. Setchell et al. aimed to determine how flares were experienced from the perspective of individuals who retrospectively characterized their flare experiences.[1] Content analyses found that increased pain was the feature most commonly associated with flares (79% of participants), followed by “other sensations” such as paresthesias, stiffness, and spasms (58%), functional limitations (50%), and changes in emotions and thinking (37%).[15] These results led to an *interim multidimensional flare definition* incorporating not only pain intensity but several other domains associated with flares: “A flare is an increase in pain or other related symptoms that lasts from hours to weeks and is difficult to settle. You may also have mood changes and/or difficulty with your normal activity.”[1] The idea that a multidimensional flare definition may capture something different from increased pain intensity was supported by data from a separate cohort of people with LBP, in which 68% of flares defined using a multidimensional flare definition were not accompanied by an increase in LBP intensity of ≥2 numeric rating scale (NRS) points, and 36% of periods with an increase in LBP intensity of ≥2 NRS points were not identified as a multidimensional flares.[4] On the other hand, this lack of complete overlap between two different flare definitions may simply reflect psychometric characteristics such as the lack of complete test-retest reliability with any self-report measure,[22] variation between any two measures evaluating the same pain construct,[21] the use of a cutoff for increased pain intensity that was quite high (≥2 points), and/or the likelihood that requiring a LBP intensity increase of ≥2 NRS points will often (but not always) select a subset of flares that are more severe.

After input and refinement from LBP experts, a Delphi process to reach consensus, and testing in a predominantly Australian sample of individuals with LBP, changes were made to the interim multidimensional flare definition: the term “worsening” was added in order to cover all potential symptoms associated with LBP rather than highlighting pain intensity alone, and the phrase “your condition” was added in order to refer not only to LBP but to related symptoms such as stiffness, spasms, and others. The resultant *final multidimensional flare definition* after these stages of input and refinement defined a flare as “…a worsening of your condition that lasts from hours to weeks that is difficult to tolerate and generally impacts your usual activities and/or emotions.”[1] This multidimensional flare definition has the strength of having involved an expert consensus process in its development, but— unlike the interim flare definition[4]— it has not yet received quantitative validation.

### 2.2 Rationale for a third approach: a person-dependent flare definition

The person-dependent flare definition derived for use in the current study was developed to (1) incorporate common elements among prior flare definitions (Table 1), and (2) address potential strengths and limitations of pain intensity-focused flare definitions and multidimensional flare definitions.

A common element across prior flare definitions is the specification of a minimum and maximum period of time during which a change in pain characteristics would be considered a flare, so that periods of worsened symptoms that are too short (e.g., a momentary increase in pain) or too long (e.g., a 6-month period of increased pain, which would be considered a new baseline level of more severe pain) would not count as a flare. Durations of “hours” to “weeks” were most commonly specified across prior flare definitions, and these bounds for duration were therefore incorporated into the current study’s person-dependent flare definition.[1]

A potential strength of the multidimensional flare definitions developed by Costa and colleagues was their expansion of the flare definition to include other pain-associated domains besides pain intensity.[1] However, there are also possible limitations from this decision. First, the qualitative methods of Setchell et al. used to inform the multidimensional flare definition were intended to capture the patient experience of flares, without a need to discriminate *components* of flares from *consequences* of flares. While the distinctions between these aspects may be immaterial to patients, combining them into a single flare definition could pose problems for researchers who wish to disentangle the components of flares from the consequences of flares. For example, if a risk factor was found to be associated with a multidimensional definition of flares, including changes in pain intensity, mood, and/or functional limitations, it would be unknown which of these three domains was driving the risk factor-flare association (Figure 1B). A second potential limitation is that the flare-associated domains included in the multidimensional flare definitions by Costa and colleagues might have been specific to the internet-based sample of Australian people in which they were derived. This could limit the definition’s external validity when applied to people from other countries, health care systems, and/or sociocultural backgrounds.

A potential strength of pain intensity-focused flare definitions is that they may allow researchers to better separate the components of flares from the consequences of flares (Figure 1A). A potential limitation of such a definition, however, is suggested by the qualitative study by Setchell et al., in which 79% of individuals with LBP perceived increased pain intensity to be a component of flares, yet 21% did not.[15],

Given these considerations, the current study evaluated the psychometric properties of a novel flare measure based on a *person-dependent flare definition* (i.e., “…a worsening of your low back pain that lasts from hours to weeks”). This definition inquires about “worsening” of LBP in an open-ended manner. When a flare is reported using a person-dependent flare definition, it is meant to be followed by *descriptive questions* that inquire specifically about the pain-related domains that are perceived to be components of the flare for a given participant. While the person-dependent flare definition is expected to include pain intensity, which is the most prevalent component of flares, a strength of the definition is that it allows for different participant perspectives on the pain-related domains that comprise flares in different populations, without the researcher specifying what those domains must be. This addresses the concern about generalizability of the multidimensional flare definitions by Costa and colleagues noted above, as flare components identified by one population are not be assumed to be applicable to another population. Another strength of the person-dependent flare definition is that, when followed by the above-mentioned descriptive questions, the descriptive questions allow the researcher to know precisely which pain domains are described as components of flares by a given participant. This allows the researcher to conduct *post hoc* analyses examining the subset of flares reflecting any combination of flare components specified by the researcher. For example, a researcher using the person-dependent flare definition could conduct secondary analyses using an alternate flare definition that emulates the final multidimensional flare definition of Costa et al. by specifying the type of flare to be analyzed as one that meets the person-dependent flare definition, but also requiring that the flare be described by the participant as difficult to tolerate, *and* either impacting usual activities *or* emotions. Alternatively, if a researcher wishes to narrow down to a more exclusively pain intensity-focused definition of flares (Figure 1A) to mitigate the potential conflation of flare components and flare consequences in a single flare outcome, they could specify an alternative flare definition that requires flares to meet the person-dependent flare definition, but also requires that flares be described by the participant as involving greater pain intensity than is usual for a participant, or having higher pain NRS ratings as compared to a participant’s non-flare periods.

### 2.3 Study sample

We evaluated the psychometric properties of a novel flare measure based on a person-dependent flare definition in an ancillary study to a longitudinal case-crossover study of people with LBP. Participants were recruited between April 2020 and September 2023 from adults seen for LBP in the Veterans Affairs Puget Sound Health Care System (VAPSHCS). VAPSHCS provides health care to military Veterans in the US states of Alaska, Idaho, Montana, Oregon, and Washington. The study was approved by the VAPSHCS Institutional Review Board (01860). Inclusion criteria included: (1) age 18 to 65 years; (2) having pain in the area of the low back between the lowest ribs and the gluteal fold, with or without lower extremity radiating symptoms; (3) regular access to a personal computer, tablet, or smartphone with internet access; (4) having a mobile phone capable of receiving text messages; (5) basic computer literacy; and (6) being able to understand and read English sufficient to provide informed consent and complete the study assessments. Exclusion criteria included: (1) pregnancy; (2) being imprisoned; (3) having a “red flag” condition such as spinal infection, malignancy, fracture, or spondyloarthropathy; (4) unstable or severe psychiatric conditions (e.g., delirium, mania, psychosis, thought disorder); (5) dementia or other significant cognitive dysfunction; (6) thoracolumbar spine surgery in the past 1 year; and (6) major orthopedic, abdominal, or chest surgery within the past 6 months. The parent study was designed primarily to examine the short- and long-term effects of physical activities on flares of LBP and was not designed specifically or exclusively for the current analyses. While the parent study included only patients seen in primary care clinics for LBP during the recruitment period (the “original cohort”), this ancillary study also analyzed an additional group of patients who had previously been seen in primary care clinics for LBP yet reported ongoing LBP (the “expanded cohort”, n=48).

### 2.4 Data collection

All data collection after enrollment used electronic, internet-based surveys, referred to as “e-Surveys” [20]. After baseline e-Surveys were completed, participants completed two types of recurring e-Surveys: (1) “Scheduled e-Surveys” and (2) “Flare Window e-Surveys”. Scheduled e-Surveys were administered 3 times per week for weeks 1-4 of follow-up, once per week for weeks 5-8 of follow-up, and twice per month thereafter during months 3-12. Flare Window e-Surveys were participant-initiated surveys available for participants to complete on an as-needed basis whenever they experienced an LBP flare, at any time during the 1-year follow-up period. Scheduled and Flare Window e-Surveys consisted of the same general content [20]. The analyses conducted to address the study aims used all available data collected until August 2024, including up to a 1-year maximum duration of follow-up for participants.

### 2.5 Participant education on the flare definition at study baseline

At the time of recruitment, participants were provided with the person-dependent flare definition used in this study (“*A worsening of your low back pain that lasts from hours to weeks*”) and received an educational session regarding this definition. Participants were advised not to consider brief increases in low back pain (lasting only seconds to minutes) as a flare, as such changes are not sustained. They were also advised not to consider increases in back pain lasting longer than one month as a flare, as a change of this type would represent a new higher baseline level of LBP rather than a flare. Participants were provided with a “Key Points” hard copy document to reinforce verbal instructions. This document specified the region where low back pain can occur using a figure indicating the area between the lowest ribs and the gluteal fold. The Key Points document also included the following written information, reiterating and clarifying the person-dependent flare definition: “A ‘flare’ is *a worsening of your low back pain that lasts from hours to weeks*, *but no longer than a month. A slight change in your low back pain intensity does not count as a flare. A worsening of your low back pain that lasts seconds to minutes does not count as a flare, because it is too short. A worsening of your low back pain that lasts 1 month or more also does not count as a flare, because it is too long*”. Participants were given the opportunity to ask any questions they had about self-identification of flares and to have questions answered by research staff. If participants had difficulty understanding the study’s flare definition at this point in the education sessions or requested more specificity on how to identify flares, the following content was also provided: *“Think of your usual (average) low back pain over the past few weeks on a scale going from 0 to 10, where 0 is* ‘*No low back pain,’ and 10 is* ‘*The worst possible low back pain that you can imagine.’ For many people, an increase in low back pain intensity of 2 or more points on this scale over their usual level of pain that lasts for hours or even weeks, might be considered a flare”*.

### 2.6 Baseline characteristics

At study baseline, participants reported on demographics and clinical factors using question items from the NIH Low Back Pain Task Force Minimal Dataset.[5] These included age; self-identified gender (man, woman, non-binary); self-identified race (American Indian or Alaska Native, Asian, Black or African-American, Native Hawaiian or Other Pacific Islander, White, or Unknown; participants were able to select one or more race categories); self-identified ethnicity (Hispanic or Latino); highest level of education attained; household income; height and weight (used to calculate body mass index); duration of LBP, and frequency of LBP (<half the days in the past 6 months vs. at least half the days vs. every day in past 6 months) using question items from the National Institutes of Health (NIH) minimum dataset for LBP.[5] LBP-related functional limitations were evaluated using the 24-item Roland-Morris Disability Questionnaire (RMDQ). The RMDQ has a score range from 0 to 24, with higher scores indicating greater pain-related disability.[14]

### 2.7 Repeated assessment of flare status and associated factors over 1-year follow-up

At each Scheduled and Flare Window e-Survey, participants were again provided with the flare definition used in the study: “*A ‘flare’** of low back pain is a worsening of your low back pain that lasts from hours to weeks.”* Additional information reiterating points from the initial education sessions was shown to participants if they sought clarification on how to identify flares by hovering their cursor over the “**” symbol in the flare definition provided. Periods when a participant reported a flare that was preceded by a period when a participant had not reported a flare were classified as a “new flare” period. Periods when a participant reported a flare that was preceded by a flare period were also classified as a “new flare” period if participants also reported that their current flare period was a new and distinct flare, above and beyond their previously-reported flare period. Alternatively, flare periods preceded by another flare period *without* their current period representing a new and distinct flare above and beyond their previous flare were classified as “flare continuation” periods. Other periods were classified as non-flare periods.

If a participant reported having a flare in a given survey, subsequent *descriptive question* items (see Section 2.2) inquired further about the flare’s characteristics. Participants reporting flares were asked, “Has your current flare involved any of the following?” and were able to choose one or more of 5 response options, each reflecting a pain-related domain from among those that had been included in the final multidimensional flare definition by Costa and colleagues[1]. These included “It has been difficult to tolerate” (which we refer to here as the *coping domain*), “It has impacted your usual activities” (the functional limitations domain), and “It has impacted your mood or emotions” (the *mood/emotions domain*). We also included two additional pain-related domains related to the theme of “increase in interventions”, such as medications or other pain treatments, based on the qualitative study by Setchell and colleagues [15] and the concept of being “difficult to settle” from the interim multidimensional flare definition by Costa and colleagues[1]: “It required more pain medication or a new pain medication to manage” (the *requiring medications domain*), and/or “It required a specific treatment to manage (other than medication)” (the *requiring other treatment* domain).[1; 4] Participants were also asked to rate the average intensity of their current LBP flare, as compared to their usual LBP intensity, choosing only one from the response options of “The same,” “A little worse,” “Much worse,” or “Very much worse”.

### 2.8 Repeated assessment of the study outcomes over 1-year follow-up

The primary study outcome was the current LBP intensity NRS. Secondary outcomes including the pain interference NRS and self-reported analgesic use. At each Scheduled and Flare Window e-Survey participants rated their current LBP intensity using a 0 to 10 NRS with the following wording: “How would you rate your low back pain intensity **<u>right now</u>**?”. Participants selected ratings of pain intensity between 0 and 10, where 0 indicated “No pain” and 10 indicated the “Worst imaginable pain”.[5] Additionally, participants reported current LBP-related pain interference using the “PEG” questionnaire pain interference item,[10] modified to refer to the low back region specifically, as follows: “What number best describes how much your low back pain interferes with your general activity, right now”. Participants selected 0 to 10 NRS ratings of pain interference between 0 and 10, where 0 indicated “Does not interfere” and 10 indicated “Completely interferes”. Participants also indicated whether they had taken pain medications (analgesics) in the past 8 hours, selecting from a list of common pain medications.[16]

### 2.9 Statistical analysis

First, we characterized the study sample according to participants’ baseline characteristics. We then conducted descriptive analyses to summarize flare frequency as the number of flares reported by participants per quarter (each 90-day period) across all Scheduled and Flare Window surveys. We examined the distribution of LBP intensity NRS ratings during flare periods and during non-flare periods. Among periods when participants reported flares, we examined the frequency of endorsement of each pain-related domain included in multidimensional flare definitions (coping, functional limitations, mood/emotions, requiring medications, requiring other treatments). We also calculated the frequency of current LBP flare ratings (“The same”, “A little worse”, “Much worse”, “Very much worse”) among periods when participants reported flares.

Next, to address the main study aim, we examined associations between flare status and pain outcomes using mixed effects linear regression models to account for repeated measures by participants. In the basic formulation of these analyses, we examined flare status as the predictor (independent) variable and current LBP intensity NRS as the primary outcome (dependent) variable, with a fixed effect intercept reflecting mean LBP intensity NRS in the absence of a flare, a fixed effect for flare status, a random intercept reflecting variation in LBP intensity NRS across persons in the absence of a flare, and a random slope reflecting variation in the flare effect on LBP intensity NRS. The primary statistical comparison of interest was whether flare status was significantly associated with current LBP intensity NRS, which was considered hypothesis testing for validity using the terminology of the Consensus-based Standards for the selection of health status Measurement Instruments (COSMIN) checklist.[11] The primary analysis excluded flare continuation periods, to focus on associations at time of flare onset; secondary analyses included flare continuation periods. We conducted unadjusted analyses first, followed by analyses adjusting for all baseline characteristics including age; self-identified gender; self-identified race; self-identified Hispanic or Latino ethnicity; education; income; body mass index; duration of LBP, and frequency of LBP. We used multiple imputation by chained equations for covariate-adjusted analyses, with 30 sets of imputations and up to 10 iterations, to recover observations with missing covariate data. Sensitivity analyses were also conducted repeating the above analytic steps restricted to Scheduled e-Surveys only.

Analogous analyses were also conducted for the secondary outcome variables of LBP-related pain interference measured by the PEG pain interference item and participant-reported analgesic use.

In a secondary analysis, similar models were run to those described above for the primary outcome variable of current LBP intensity NRS (with and without adjustment for baseline covariates) using an “alternative flare definition”. This alternative flare definition was intended to emulate the final multidimensional flare definition by Costa and colleagues (Table 1),[1] requiring not only participant endorsement of a current participant-dependent flare, but also requiring endorsement that the flare was difficult to tolerate (the coping domain) and endorsement of *either* of the mood/emotions or functional limitations domains.

## 3. RESULTS

### 3.1. Characteristics of the study sample

Four hundred sixty-five Veterans were consented and comprised the study sample. The mean ± standard deviation (SD) age of the study sample was 48 ± 11 years, and 26% of participants were women (Table 2), reflective of the US military Veteran population, which is comprised predominantly of men. The majority of participants (75%) self-reported their racial group as White. Seventeen percent identified as being Black/African-American, 7% identified as being Asian, 7% identified as being American Indian/Alaska Native, 2% identified as being Native Hawaiian/Pacific Islander, and 9% identified as being from more than one racial group. Twelve percent of the sample reported being of Latino or Hispanic ethnicity. About half of the sample had a college degree (48%), and 40% had an average household income more than the US median ($75,000).[8] Nearly all participants (96%) reported LBP that had been present for more than 3 months (the most common duration used to define LBP as “chronic”[5]), and 78% reported their LBP had been present for ≥ 5 years.

**Table 2.**
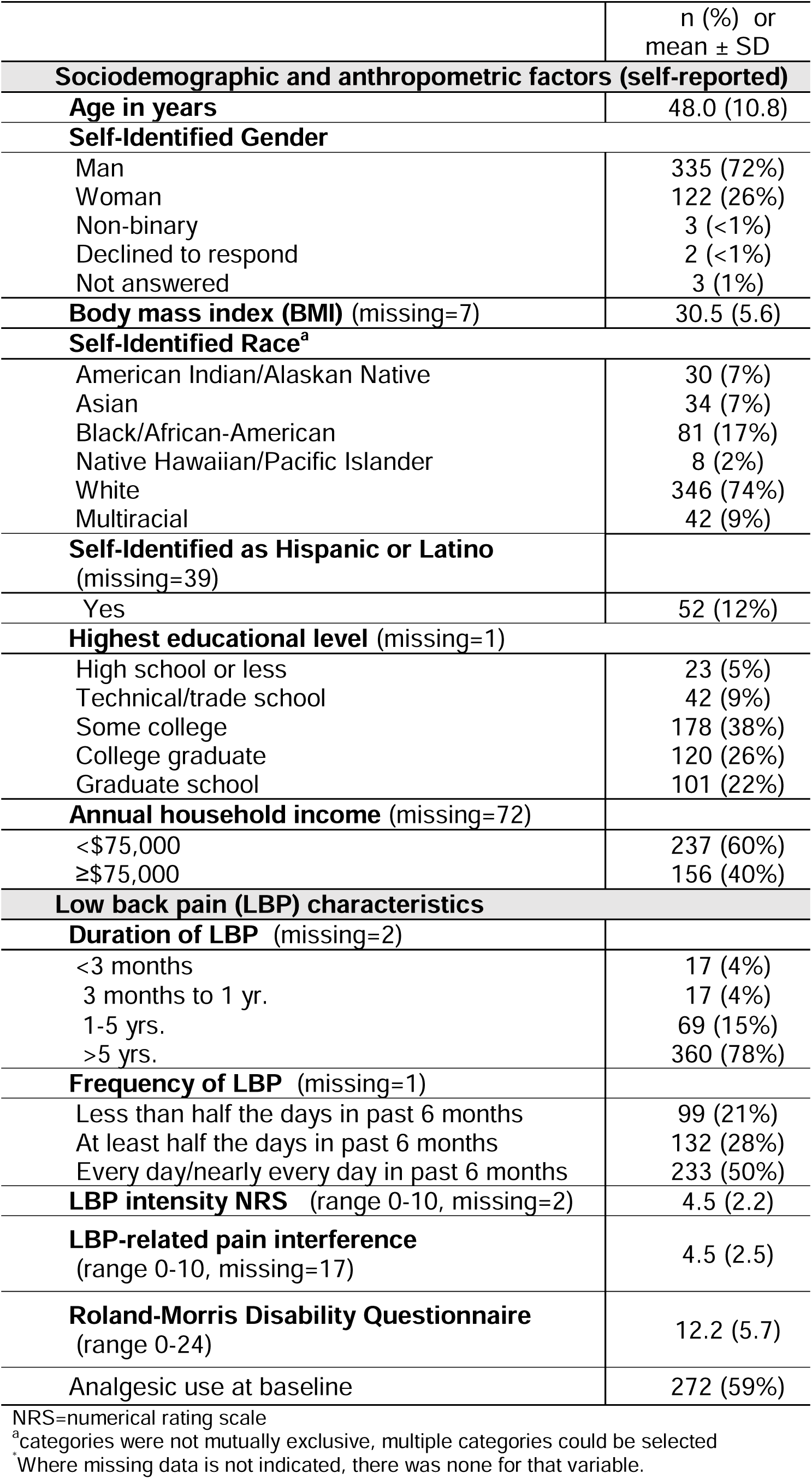
Baseline characteristics of the Study Sample (n=465)*

| **Table 2. Baseline characteristics of the Study Sample (n=465)\*** |  |
| --- | --- |
| | n (%) or<br>mean ± SD |
| **Sociodemographic and anthropometric factors (self-reported)** |  |
| **Age in years** | 48.0 (10.8) |
| **Self-Identified Gender** |  |
| Man | 335 (72%) |
| Woman | 122 (26%) |
| Non-binary | 3 (<1%) |
| Declined to respond | 2 (<1%) |
| Not answered | 3 (1%) |
| **Body mass index (BMI) (missing=7)** | 30.5 (5.6) |
| **Self-Identified Race<sup>a</sup>** |  |
| American Indian/Alaskan Native | 30 (7%) |
| Asian | 34 (7%) |
| Black/African-American | 81 (17%) |
| Native Hawaiian/Pacific Islander | 8 (2%) |
| White | 346 (74%) |
| Multiracial | 42 (9%) |
| **Self-Identified as Hispanic or Latino (missing=39)** |  |
| Yes | 52 (12%) |
| **Highest educational level (missing=1)** |  |
| High school or less | 23 (5%) |
| Technical/trade school | 42 (9%) |
| Some college | 178 (38%) |
| College graduate | 120 (26%) |
| Graduate school | 101 (22%) |
| **Annual household income (missing=72)** |  |
| <\$75,000 | 237 (60%) |
| ≥\$75,000 | 156 (40%) |
| **Low back pain (LBP) characteristics** |  |
| **Duration of LBP (missing=2)** |  |
| <3 months | 17 (4%) |
| 3 months to 1 yr. | 17 (4%) |
| 1-5 yrs. | 69 (15%) |
| >5 yrs. | 360 (78%) |
| **Frequency of LBP (missing=1)** |  |
| Less than half the days in past 6 months | 99 (21%) |
| At least half the days in past 6 months | 132 (28%) |
| Every day/nearly every day in past 6 months | 233 (50%) |
| **LBP intensity NRS (range 0-10, missing=2)** | 4.5 (2.2) |
| **LBP-related pain interference (range 0-10, missing=17)** | 4.5 (2.5) |
| **Roland-Morris Disability Questionnaire (range 0-24)** | 12.2 (5.7) |
| **Analgesic use at baseline** | 272 (59%) |

NRS=numerical rating scale
<sup>a</sup>categories were not mutually exclusive, multiple categories could be selected
\*Where missing data is not indicated, there was none for that variable.

### 3.2 Flare frequencies

Participants completed 11,884 surveys, of which 9891 (83%) were Scheduled e-Surveys and 1993 (17%) were Flare Window e-Surveys. Among the Scheduled e-Surveys, 78% of periods were non-flare, 16% were new flares, and 5% were flare continuation periods. Among Flare Window surveys, 88% of periods were new flares, and 12% were flare continuation periods.

The mean number of flares per quarter (90 days) was 5.1, with a standard deviation (SD) of 9.2. The distribution of the number of flares per quarter is provided in Supplemental Figure 1, which shows that most participants (65%) reported 0 to 4 flares per quarter.

### 3.3. LBP NRS scores by flare status

Histograms of LBP intensity NRS scores are provided in Supplemental Figures 2-3. Pain NRS scores during participants’ first flare periods were roughly normally distributed, with a median NRS of 6.0. Pain NRS scores during participants’ first non-flare periods were right-skewed, with a median NRS of 3.0. Similar distributions of NRS scores and patterns of skewness by flare status were observed when participants’ worst flare and non-flare periods were analyzed, and when analyses were restricted to Scheduled e-Surveys alone (data not shown).

### 3.4 Description of flare status by participant perceptions of changes in pain-related domains and pain intensity

During periods when participants reported new flares (n=3382), participants reported that their flares were associated with impacts on coping (45% of periods), functional limitations (68%), mood/emotions (57%), requiring analgesics to manage (34%), and requiring another treatment (i.e., something other than an analgesic) to manage (13%) (Table 3). During nearly all periods when participants reported flares, impacts were reported in at least one pain-related domain, and most periods were reported to have impacted 1 or 2 pain-related domains (Table 3). The prevalences of reported impacts on each pain-related domain and the number of pain-related domains were consistently higher during flare continuation periods than during new flare periods.

**Table 3.** Prevalence of ratings of pain-related domains and other factors, by flare status, during periods when flares were reported*.

|  | New flares<br>(n=3382 periods) | Flare continuation<br>periods (n=751 periods) | Any flare (new or<br>continuation, n=4133<br>periods) |
| --- | --- | --- | --- |
| **Participant-reported impact on pain-related domains** |  |  |  |
| Coping | 1503 (45%) | 393 (52%) | 1896 (46%) |
| Functional limitations | 2258 (68%) | 570 (76%) | 2829 (69%) |
| Mood | 1926 (57%) | 481 (64%) | 2407 (58%) |
| Requiring analgesics | 1160 (34%) | 340 (45%) | 1500 (36%) |
| Requiring other treatment | 436 (13%) | 99 (13%) | 535 (13%) |
| **Number of pain-related domains impacted** |  |  |  |
| No domains impacted | 3 (<1%) | 0 | 3 (<1%) |
| 1 domain | 1189 (36%) | 231 (31%) | 1420 (35%) |
| 2 domains | 797 (24%) | 146 (19%) | 943 (23%) |
| 3 domains | 783 (23%) | 134 (18%) | 917 (22%) |
| 4 domains | 430 (13%) | 194 (26%) | 624 (15%) |
| 5 domains | 140 (4%) | 46 (6%) | 186 (5%) |
| **Participant descriptions of current LBP intensity (during flare) as compared to usual LBP intensity (without flare)<sup>a</sup>** |  |  |  |
| "the same" | 546 (17%) | 198 (27%) | 744 (19%) |
| "a little worse" | 1373 (42%) | 280 (38%) | 1653 (41%) |
| "much worse" | 1162 (35%) | 211 (29%) | 1373 (34%) |
| "very much worse" | 212 (6%) | 47 (6%) | 259 (6%) |

\*All variables are participant-reported assessments
<sup>a</sup>"How would you rate the average intensity of your current low back pain flare, compared to your usual low back pain intensity?"

When participants reporting flares rated their perceptions of current LBP intensity (during a flare) as compared to usual LBP intensity (without flare), most participants reporting new flares reported their current LBP intensity was “a little worse” (42% of periods) or “much worse” (35% of periods) than their usual LBP intensity when not having a flare (Table 3). However, during 17% of periods when participants reported new flares, participants reported their current LBP intensity during the flare was “the same” as their usual LBP intensity.

### 3.5 Associations between flare status and LBP intensity: hypothesis testing

Pain NRS ratings were higher during new flare periods (median NRS=6) and flare continuation periods (median NRS=7) than non-flare periods (median NRS=2) and the distribution of non-flare periods was right-skewed due to a greater frequency of lower pain ratings (Supplemental Figure 4). Subsequent analyses examining the associations between participant-reported flare status and pain intensity analyzed only new flare and non-flare periods (excluding flare continuation periods), as had been pre-specified.

In an unadjusted linear mixed-effect model examining associations between flare status and current LBP intensity NRS, the primary outcome variable for the study aim, flare periods were highly significantly associated with a 2.8-NRS point greater pain intensity fixed effect as compared to non-flare periods (95% CI 2.7-2.9, *p*<0.0001) in unadjusted analyses (Table 4), with a 1.2-NRS point standard deviation of the flare “effect.” The fixed effect intercept was 2.8, indicating an average LBP intensity of 2.8 NRS points during periods in which participants were <u>not</u> having a flare, with a standard deviation of 1.9 NRS points. In multivariable analyses adjusting for all baseline variables from Table 1, the association of flare status with LBP intensity NRS was unchanged (Table 4), with a 2.8-NRS point greater LBP intensity NRS fixed effect as compared to non-flare periods (95% CI 2.7-3.0, *p*<0.0001).

**Table 4.** Hypothesis testing: associations between flare status during new flare periods and reference standard pain measures.

|  | **Fixed effects** |  | **Random effects** |  |
| --- | --- | --- | --- | --- |
| Model estimates | Flare status | Model intercept | Flare status standard deviation (SD) | Model intercept standard deviation (SD) |
| Interpretation | Difference in LBP intensity NRS (95% CI) with flare periods compared to non-flare periods | Average LBP intensity NRS (95% CI) during non-flare periods | Variability of LBP intensity NRS response to flare | Variability of participants' LBP intensity NRS during non-flare periods |
| Unadjusted model | 2.8 (2.7-2.9) | 2.8 (2.7-3.0) | 1.2 | 1.9 |
| Adjusted model<sup>a</sup> | 2.8 (2.6-2.9) | 1.4 (0.2-2.6) | 1.1 | 1.6 |

**Associations between flare status and LBP-related pain interference NRS (secondary outcome)**
|  | **Fixed effects** |  | **Random effects** |  |
| --- | --- | --- | --- | --- |
| Model estimates | Flare status | Model intercept | Flare status standard deviation (SD) | Model intercept standard deviation (SD) |
| Interpretation | Difference in pain interference (95% CI) with flare periods compared to non-flare periods | Average pain interference (95% CI) during non-flare periods | Variability of pain interference response to flare | Variability of participants' pain interference during non-flare periods |
| Unadjusted model | 2.9 (2.8-3.1) | 2.7 (2.5-2.9) | 1.4 | 2.1 |
| Adjusted model<sup>a</sup> | 2.9 (2.7-3.1) | 2.3 (1.0-3.6) | 1.4 | 1.7 |

**Associations between flare status and analgesic use (secondary outcome)**
|  | **Fixed effects** |  | **Random effects** |
| --- | --- | --- | --- |
| Model estimates | Flare status |  | Flare status standard deviation (SD) |
| Interpretation | Odds ratio for analgesic use (95% CI) with flare periods compared to non-flare periods |  | Variability of analgesic use response to flare<sup>b</sup> |
| Unadjusted model | 4.1 (3.3-5.2) |  | 1.3 |
| Adjusted model<sup>c</sup> | 4.0 (3.3-5.2) |  | 1.3 |

LBP=low back pain, NRS=numeric rating scale, CI=confidence interval
<sup>a</sup>Adjusted models included covariates of age; self-identified gender, self-identified race, self-identified Hispanic or Latino ethnicity, education, income, body mass index, duration of LBP, and frequency of LBP.
<sup>b</sup>log-odds scale
<sup>c</sup>Adjusted models included only a subset of covariates due to problems with model convergence: self-identified gender, body mass index, and education.

### 3.6 Associations between flare status and LBP-related pain interference and analgesic use: hypothesis testing

In an unadjusted mixed-effect linear model examining associations between flare status and LBP-related pain interference NRS, flare periods were significantly associated with a 2.9-NRS point greater LBP-related pain interference fixed effect as compared to non-flare periods (95% CI 2.8-3.1, *p*<0.0001, Table 4), with a 1.4-NRS point standard deviation of the flare “effect”. The fixed effect intercept was 2.7, indicating an average LBP-related pain interference of 2.7 NRS points during periods in which participants were <u>not</u> having a flare, with a standard deviation of 1.7 NRS points. In multivariable analyses adjusting for all baseline variables from Table 1, the association of flare status with LBP-related pain interference was unchanged (Table 4), with a 2.9-NRS point greater pain interference fixed effect as compared to non-flare periods (95% CI 2.7-3.1, *p*<0.0001).

In mixed-effects linear models examining associations between flare status and analgesic use, flare periods were highly significantly associated with a 4-fold increase in the log odds of analgesic use as compared to non-flare periods (odds ratio [OR]=4.1, 95% CI 3.3-5.2; *p*<0.0001) in unadjusted analyses (Table 3). In multivariable analyses adjusting for all baseline variables from Table 2, the association of flare status with LBP-related pain interference was not materially different (OR=4.0, *p*<0.0001; Table 4).

### 3.7 Associations between flare status and the alternative flare definition

Of 3,382 new flare periods reported using the person-dependent flare definition, 1,314 (39%) also met criteria for the alternate flare definition proxying the final multidimensional flare definition by Costa and colleagues. In mixed-effects linear models examining associations between the alternative flare definition and LBP-related pain interference NRS reflected by the PEG pain interference question item, flare periods were statistically significantly associated with a 2.8-NRS point greater LBP-related pain intensity fixed effect as compared to non-flare periods in both unadjusted (95% CI 2.6-2.9, *p*<0.0001) and adjusted analyses (95% CI 2.6-3.90, *p*<0.0001), highly consistent with the primary analyses of the person-dependent flare definition

### 3.8 Analyses to understand flare periods when participants reported that their LBP intensity was the same as their usual LBP intensity

We conducted *post hoc* analyses to further examine the finding that 19% of flare periods (17% of new flares and 27% of flare continuation periods) were described by participants as having “the same” LBP intensity as their usual LBP when not in flare. In the subset of flare periods reported as having “the same” LBP intensity as usual, 28% of periods were reported as having an impact on coping. In comparison, the corresponding proportions were 52% for impact on functional limitations, 46% for impact on mood, and relatively lower proportions for the domains of requiring analgesics (19%) and requiring other treatments (9%) (Supplemental Table S1). Participant impressions of greater severity of their LBP intensity during flare vs. non-flare periods were associated with greater perceived impact on the five pain-related domains of coping, functional limitations, mood/emotions, requiring analgesics, and requiring other treatments (Supplemental Table S1). Among periods when participants who reported flares also reported that their current LBP intensity was the same as their usual back pain intensity, 57% were associated with participant perceptions (in the past 24 hours) of having had brief increases in LBP intensity not sufficient to count as flares by themselves, and 42% with feeling like their back was weaker or more vulnerable than usual.

In *post hoc* analyses to describe within-person change in LBP intensity, we calculated the frequency of flare periods during which a person’s LBP intensity was less than 2 NRS points greater than the same person’s average pain intensity during non-flare periods. Thirty-seven percent of flare periods had LBP intensity values <2 NRS points compared to their average pain intensity during non-flare periods. When using mixed-effects linear regression models to characterize flare period-pain intensity associations *within* strata of perceived LBP intensity increase during flare periods, flares perceived as having “the same” LBP intensity as usual were associated with a 2.0-NRS-point greater average LBP intensity when compared to non-flare periods, a value lower than the 2.8-NRS point greater average in the whole sample, but one that nevertheless indicated a substantial increase over usual pain intensity. (Supplemental Table S2). In higher strata of perceived increase in LBP intensity as compared to usual LBP intensity, estimates of flare-LBP intensity associations were progressively larger (Supplemental Table S2).

## 4. DISCUSSION

This longitudinal study was the largest ever to validate a definition of “flare” among people with LBP. The study’s main finding was that LBP flares measured based on a participant-dependent flare definition (i.e., “A flare of low back pain is a worsening of your low back pain that lasts from hours to weeks”) were highly significantly associated with more severe pain outcomes on validated LBP-related outcome measures, with or without adjustment for person-level factors. Participant flare periods were associated with greater LBP intensity NRS values than non-flare periods by an average of nearly 3 NRS points, a large difference well above the commonly accepted minimal clinically important change (MCIC) values of 1-2 NRS points.[12] Similarly, participant flare periods were highly significantly associated with greater LBP pain interference by nearly 3 NRS points and a 4-fold increase in the odds of analgesic use, as compared to non-flare periods. New flare periods were perceived as having impacts on many pain-related domains (e.g., coping, functional limitations, mood/emotions, and others). These results provide strong support for the convergent validity of the measure based on this participant-dependent flare definition.

The findings of the current study comport with prior studies examining the convergent validity of flare definitions with pain intensity. In the first longitudinal study to validate a flare definition, Suri and colleagues found that a pain intensity-focused flare definition (i.e., “…a period of increased pain lasting at least 2 hours, when your pain intensity is distinctly worse than it has been recently”) was significantly associated with a 2.7-NRS-point greater pain intensity during flare vs. non-flare periods, nearly the same magnitude association with flare status (2.8 NRS points) found in the current study.[17] Similarly, Costa and colleagues reported statistically significant greater average LBP intensity during flare vs. non-flare periods of between 1.3 to 3.3 NRS points when assessing the presence of an LBP flare using a pain intensity-focused definition, and smaller-magnitude yet still significant differences in LBP intensity between flare vs. non-flare periods (between 0.3 and 0.9 NRS points) when a multidimensional pain definition was used. In addition, and also similar to the current study’s finding of significantly greater pain interference and analgesic use during flare periods vs. non-flare periods, Costa and colleagues found greater LBP-related pain interference and analgesic use with both pain intensity-focused and multidimensional flare definitions, although pain intensity-focused definitions generally had larger-magnitude associations with flare status than did multidimensional flare definitions for non-pain intensity outcomes such as LBP-related pain interference, mood-related outcomes, and most other outcomes (with the exception of analgesic use).[4] The findings of the current study are generally consistent with results derived from qualitative research that flares involve dimensions that are broader than pain intensity alone.[15] On the other hand, one could argue that the experience of pain in general impacts dimensions and domains broader than pain intensity alone, so in this respect flares are no different than any other aspect of the pain experience. Of note, the proportion of flare periods in which participants in the current study reported the co-existence of impacts on coping, functional limitations, and mood/emotions exceeded the prevalences found in the qualitative research of Setchell and colleagues,[15] which may be explained by the different study designs, settings, and flare definitions used.

During 19% of flare periods in the current study, participants reported their LBP intensity was roughly the same as their usual LBP when not having flare, indicating that respondent interpretations of the study’s participant-dependent flare definition did not require perceived “worsening” to be defined by increases in pain intensity alone. This proportion closely resembles the finding of Setchell and colleagues that 21% of people with flares did not report pain intensity increase as part of the flare experience.[15] In the current study, some proportion of flare periods reported as having “the same” LBP intensity as usual were reported to impact coping (28%) or mood/emotions (46%), such that pain intensity may have remained the same, but participants’ ability to cope with that pain may have decreased, or they may have developed mood impairments (e.g., greater depression symptoms) as a result of being in pain, resulting in the perception of a flare taking place. Additionally, large proportions of flare periods were reported as having the same LBP intensity as usual were associated with brief, transient increases in pain (57%) or feelings like the back was weaker or more vulnerable than usual (42%) in the preceding 24 hours, either of which might have contributed to the perception of a flare even when current LBP intensity was no different during flare vs. non-flare periods. However, another explanation for the 19% of flare periods reported as having roughly the same LBP intensity as usual could simply be imprecision or inaccuracy in reporting caused by the subjective nature of pain assessments. Such an explanation is suggested by *post hoc* analyses indicating that even during flare periods when participants reported that their LBP intensity was the same as usual, their average LBP intensity was 2 NRS points higher than during non-flare periods.

A strength of this study is the large number of participants and very large number of periods analyzed (>11,000) over 1-year of follow-up. Another unique strength is that, as reported elsewhere,[19] the current study sample has been shown to be representative of the underlying study population of military Veterans with LBP served by the VAPSHCS healthcare system serving 5 states in the western US, with respect to a wide range of sociodemographic, medical, psychological, and other clinical factors. A study limitation is that although we established the convergent validity of the study’s participant-dependent flare definition as compared to LBP intensity, LBP-related pain interference, and analgesic use, we did not conduct similar analyses for other important pain-related domains (e.g., changes in depression, coping, etc.). Further, one could argue that the participant-dependent flare definition for LBP used in the current study implies pain intensity as a requisite for flare, as participants who had difficulty understanding the study’s flare definition and required more information to distinguish flare vs. non-flare periods were encouraged to consider an increase in their LBP intensity ≥2 NRS points over their usual LBP intensity as a tool to help them identify flares. Nevertheless, the current study’s finding that 19% of participants reported their LBP intensity during flare was the same as their usual LBP intensity argues against the idea that participants believed that an increase in LBP intensity over their usual intensity is always necessary for such an event to “count” as a flare. Moreover, prior work using multidimensional flare definitions that included more explicit specification of increased pain intensity as a requirement for flare (“an increase in pain…”) have concluded that flares using such definitions were not defined by pain intensity alone.[4] Finally, the current study does not examine whether participant-dependent flare definitions are better or worse than pain intensity-focused or multidimensional pain definitions, which is a worthwhile future research direction.

Despite possible limitations, the current study’s findings support the convergent validity of a new measure of LBP flares based on a participant-dependent flare definition. This participant-dependent flare definition can be used by researchers to examine the frequency and impact of LBP flares, as well as the impact of interventions designed to decrease LBP flares.

## ACKNOWLEDGEMENTS

Dr. Suri, Dr. Korpak, Mr. Timmons, Ms. Tanus, Ms. Brubeck, and Dr. Daniels are employees of the VA Puget Sound Health Care System in Seattle, Washington. This research was supported by grant I01RX003248 from the Rehabilitation Research and Development Service, VA Office of Research and Development. Drs. Suri, Heagerty, and Rundell are Core Directors of the University of Washington Clinical Learning, Evidence And Research (CLEAR) Center for Musculoskeletal Research, which is funded by P30AR072572 from the National Institute of Arthritis and Musculoskeletal and Skin Diseases (NIAMS) of the National Institutes of Health. The content is solely the responsibility of the authors and does not necessarily represent the official views of the US Department of Veterans Affairs, the National Institutes of Health, or the US Government. The authors have no conflicts of interest. Dr. Hodges is supported by a fellowship from the National Health and Medical Research Council (Australia; #1194937). Dr. Staab was supported by a postdoctoral fellowship from the National Center for Complementary and Integrative Health (NCCIH), funded by T90 AT008544.

## DATA AVAILABILITY

The study’s informed consent processes do not allow study datasets containing individual-level data to be made available outside the federal government.

## REFERENCES

[1] Costa N, Ferreira ML, Setchell J, Makovey J, Dekroo T, Downie A, Diwan A, Koes B, Natvig B, Vicenzino B, Hunter DJ, Roseen E, Rasmussen-Barr E, Guillemin F, Hartvigsen J, Bennel K, Costa L, Macedo L, Pinheiro M, Underwood M, Van Tulder M, Johansson M, Enthoven P, Kent P, O’Sullivan P, Suri P, Genevay S, PW. H. A definition of ‘flare’ in low back pain (LBP): A multiphase process involving perspectives of individuals with LBP and expert consensus. J Pain 2019;Epub ahead of print.

[2] Costa N, Ferreira ML, Cross M, Makovey J, Hodges PW. How is symptom flare defined in musculoskeletal conditions: A systematic review. Semin Arthritis Rheum 2018;48(2):302–317.

[3] Costa N, Smits EJ, Kasza J, Salomoni S, Rodriguez-Romero B, Ferreira ML, Hodges PW. Are objective measures of sleep and sedentary behaviours related to low back pain flares? Pain 2022;163(9):1829–1837.

[4] Costa N, Smits EJ, Kasza J, Salomoni SE, Ferreira M, Hodges PW. Low Back Pain Flares: How do They Differ From an Increase in Pain? Clin J Pain 2021;37(5):313–320.

[5] Deyo RA, Dworkin SF, Amtmann D, Andersson G, Borenstein D, Carragee E, Carrino J, Chou R, Cook K, Delitto A, Goertz C, Khalsa P, Loeser J, Mackey S, Panagis J, Rainville J, Tosteson T, Turk D, Von Korff M, Weiner DK. Report of the NIH Task Force on research standards for chronic low back pain. Physical therapy 2015;95(2):e1–e18.

[6] Dieleman JL, Cao J, Chapin A, Chen C, Li Z, Liu A, Horst C, Kaldjian A, Matyasz T, Scott KW, Bui AL, Campbell M, Duber HC, Dunn AC, Flaxman AD, Fitzmaurice C, Naghavi M, Sadat N, Shieh P, Squires E, Yeung K, Murray CJL. US Health Care Spending by Payer and Health Condition, 1996-2016. JAMA 2020;323(9):863–884.

[7] Global Burden of Disease 2017 Disease and Injury Incidence and Prevalence Collaborators. Global, regional, and national incidence, prevalence, and years lived with disability for 354 diseases and injuries for 195 countries and territories, 1990-2017: a systematic analysis for the Global Burden of Disease Study 2017. Lancet 2018;392(10159):1789–1858.

[8] Guzman G, Kollar M. Income in the United States: 2022. In: USC Bureau editor. https://www.census.gov/content/dam/Census/library/publications/2023/demo/p60-279.pdf, 2023.

[9] Kongsted A, Kent P, Axen I, Downie AS, Dunn KM. What have we learned from ten years of trajectory research in low back pain? BMC Musculoskelet Disord 2016;17:220.

[10] Krebs EE, Lorenz KA, Bair MJ, Damush TM, Wu J, Sutherland JM, Asch SM, Kroenke K. Development and initial validation of the PEG, a three-item scale assessing pain intensity and interference. J Gen Intern Med 2009;24(6):733–738.

[11] Mokkink LB, Terwee CB, Patrick DL, Alonso J, Stratford PW, Knol DL, Bouter LM, de Vet HC. The COSMIN checklist for assessing the methodological quality of studies on measurement properties of health status measurement instruments: an international Delphi study. Qual Life Res 2010;19(4):539–549.

[12] Ostelo RW, Deyo RA, Stratford P, Waddell G, Croft P, Von Korff M, Bouter LM, de Vet HC. Interpreting change scores for pain and functional status in low back pain: towards international consensus regarding minimal important change. Spine (Phila Pa 1976) 2008;33(1):90–94.

[13] Ricci JA, Stewart WF, Chee E, Leotta C, Foley K, Hochberg MC. Back pain exacerbations and lost productive time costs in United States workers. Spine (Phila Pa 1976) 2006;31(26):3052–3060.

[14] Roland M, Fairbank J. The Roland-Morris Disability Questionnaire and the Oswestry Disability Questionnaire. Spine (Phila Pa 1976) 2000;25(24):3115–3124.

[15] Setchell J, Costa N, Ferreira M, Makovey J, Nielsen M, Hodges PW. What constitutes back pain flare? A cross sectional survey of individuals with low back pain. Scand J Pain 2017;17:294–301.

[16] Suri P, Heagerty PJ, Timmons A, Jensen MP. Description and initial validation of a novel measure of pain intensity: the Numeric Rating Scale of Underlying Pain without concurrent Analgesic use. Pain 2024.

[17] Suri P, Rainville J, Fitzmaurice GM, Katz JN, Jamison RN, Martha J, Hartigan C, Limke J, Jouve C, Hunter DJ. Acute low back pain is marked by variability: An internet-based pilot study. BMC Musculoskelet Disord 2011;12:220.

[18] Suri P, Saunders KW, Von Korff M. Prevalence and characteristics of flare-ups of chronic nonspecific back pain in primary care: a telephone survey. Clin J Pain 2012;28(7):573–580.

[19] Suri P, Tanus AD, Stanaway IB, Scott H, Brubeck HF, Irimia B, Daniels CJ, Jensen MP, Rundell SD, Timmons AK, Morelli D, Heagerty PJ. Evaluating the representativeness of a cohort study of low back pain: using electronic health record data to make direct comparisons of study participants with the study population. Journal of Pain 2024;Submitted to journal.

[20] Suri P, Tanus AD, Torres N, Timmons A, Irimia B, Friedly JL, Korpak A, Daniels C, Morelli D, Hodges PW, Costa N, Day MA, Heagerty PJ, Jensen MP. The Flares of Low back pain with Activity Research Study (FLAReS): study protocol for a case-crossover study nested within a cohort study. BMC Musculoskeletal Disorders 2022;23(1).

[21] Swan K, Speyer R, Scharitzer M, Farneti D, Brown T, Woisard V, Cordier R. Measuring what matters in healthcare: a practical guide to psychometric principles and instrument development. Front Psychol 2023;14:1225850.

[22] Vilagut G. Encyclopedia of Quality of Life and Well-Being Research. Cham: Springer International Publishing AG, 2024.

[23] Von Korff M. Studying the natural history of back pain. Spine (Phila Pa 1976) 1994;19(18 Suppl):2041S–2046S.

