## Supplemental File Tables and Figures for "Convergent validity of a person-dependent definition of a low back pain flare"

| **Supplemental Table 1. Participant perceptions of their current LBP intensity (during flare) as compared to their usual LBP intensity (without flare), according to perceived impact on pain-related domain** | |
| --- | --- |
| **Variables evaluated with respect to the current flare and/or the current time** | |
| **Impact on coping (“**it has been difficult to tolerate**”)** | |
|  | **Yes**, impact on coping |
| “the same” (n=744) | 208 (28%) |
| “a little worse” (n=1653) | 580 (35%) |
| “much worse” (n=1373) | 849 (62%) |
| “very much worse” (n=259) | 232 (90% |
| **Impact on functional limitations (“**It has impacted your usual activities**”)** | |
| “the same” (n=744) | 386 (52%) |
| “a little worse” (n=1653) | 1051(64%) |
| “much worse” (n=1373) | 1110 (81%) |
| “very much worse” (n=259) | 236 (91%) |
| **Impact on mood/emotions (“**It has impacted your mood or emotions **”)** | |
| “the same” (n=744) | 344 (46%) |
| “a little worse” (n=1653) | 924 (56%) |
| “much worse” (n=1373) | 895 (65%) |
| “very much worse” (n=259) | 219 (85%) |
| **Requiring analgesics to manage (“**It required more pain medication or a new pain medication to manage**”)** | |
| “the same” (n=744) | 139 (19%) |
| “a little worse” (n=1653) | 517 (31%) |
| “much worse” (n=1373) | 654 (48%) |
| “very much worse” (n=259) | 166 (64%) |
| **Requiring other treatments to manage(“**It required a specific treatment to manage (other than medication)**”)** | |
| “the same” (n=744) | 66 (9%) |
| “a little worse” (n=1653) | 162 (9%) |
| “much worse” (n=1373) | 217 (16%) |
| “very much worse” (n=259) | 80 ( 31%) |
| **Variables evaluated with respect to a 24-hour recall period^a^** | |
| **“Brief increases in your low back pain intensity not sufficient to count as a flare”** | |
| “the same” (n=744) | 424 (57%) |
| “a little worse” (n=1653) | 984 (60%) |
| “much worse” (n=1373) | 842 (61%) |
| “very much worse” (n=259) | 176 (68%) |
| **“Feeling like your back is weaker or more vulnerable than usual”** | |
| “the same” (n=744) | 311 (42%) |
| “a little worse” (n=1653) | 965 (58%) |
| “much worse” (n=1373) | 945 (69%) |
| “very much worse” (n=259) | 185 (71%) |
| ^a^These question items use a 24-hour recall period prior to the time of reporting, rather than prior to flare onset. Therefore, the time frame of recall for these variables includes substantial time *after* the expected onset of flare, and therefore may partially reflect consequences of the flare itself. Accordingly, caution is suggested with interpretation of associations between this participant descriptions of how their current LBP intensity (during flare) compares to their usual LBP intensity (without flare). | |

| **Supplemental Table S2.** Associations between flare status during new flare periods and LBP intensity NRS, when stratifying by participant perceptions of how their current LBP intensity (during flare) compares to their usual LBP intensity (without flare). | |
| --- | --- |
|  | **Odds ratio (95% confidence interval) for effect of flare status on LBP intensity NRS** |
| “the same” | 2.0 (1.7-2.2) |
| “a little worse” | 2.5 (2.3-2.6) |
| “much worse” | 3.4 (3.2-3.5) |
| “very much worse” | 4.5 (4.2-4.9) |

**Supplemental Figure 1**

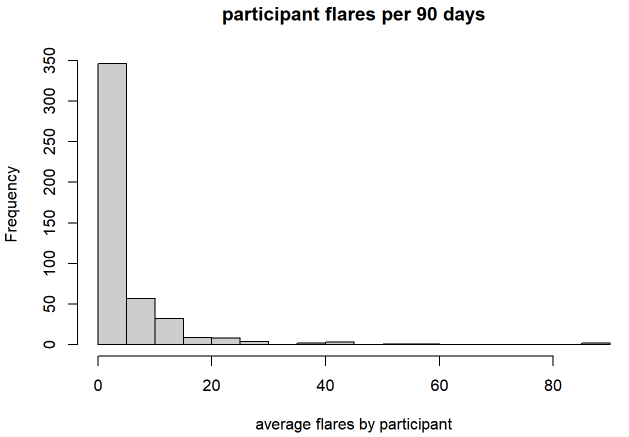
**Supplemental Figure 2**

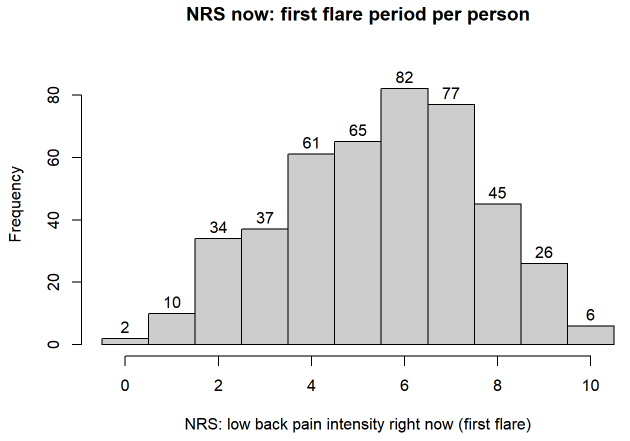

**Supplemental Figure 3**

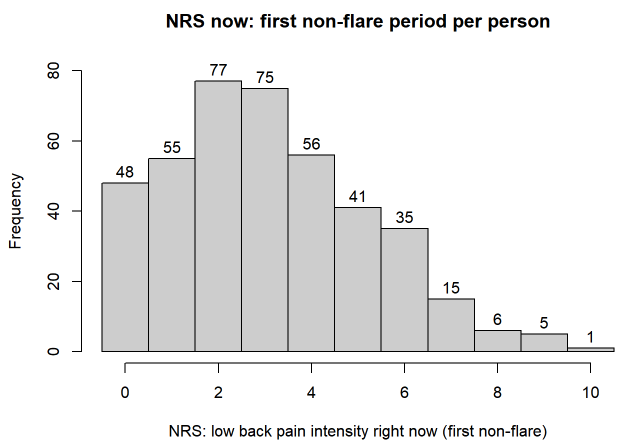

**Supplemental Figure 4**

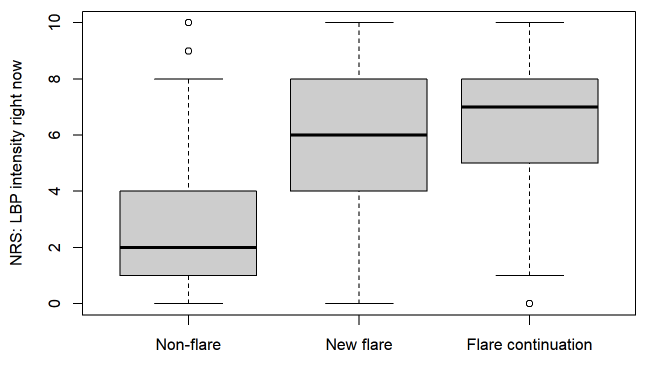
